# Neuroimmune signatures linking inflammatory proteomics to temporal cortical structure in mothers who perpetrated child maltreatment

**DOI:** 10.64898/2026.07.12.26357844

**Authors:** Sawa Kurata, Shota Nishitani, Natasha Y.S. Kawata, Akiko Yao, Ryoko Kasaba, Ryo Kuboshita, Saori Nishikawa, Takeshi Morimoto, Yasutaka Fushimi, Hidehiko Okazawa, Takashi X. Fujisawa, Akemi Tomoda

## Abstract

Neurobiological mechanisms underlying child maltreatment perpetration remain poorly understood, and the role of immune dysregulation has rarely been examined. Here, we tested whether peripheral inflammatory signatures are linked to brain structural alterations in mothers who have perpetrated maltreatment, and whether such alterations mediate this link to perpetration. In this cross-sectional study integrating structural MRI and inflammatory proteomics, 16 mothers with histories of maltreatment perpetration and 145 age-matched control mothers underwent brain imaging; a subgroup (n = 52; 11 maltreatment, 41 control) also completed plasma proteomic profiling using the Olink Target 96 Inflammation panel. Whole-brain voxel-based morphometry revealed significantly reduced gray matter volume (GMV) in the right middle/inferior temporal gyri—a region implicated in social cognition and contextual interpretation—in the maltreatment group. Proteomic analysis identified 16 inflammation-related proteins differentially expressed between groups; among these, nine were significantly associated with GMV in this temporal region. Lower GMV was associated with higher levels of pro-inflammatory proteins (CCL20, IL-17C) and with lower levels of immune-regulatory and metabolic proteins (CXCL1, CXCL6, SIRT2, STAMBP, MCP-2, MCP-4, 4E-BP1). Mediation analyses revealed that both protein sets were indirectly associated with perpetration through this regional GMV, with opposing patterns of direct association. These findings suggest that peripheral immune imbalance—characterized by elevated inflammatory signaling and diminished immune-regulatory capacity—is linked to structural vulnerability in a temporal cortical region involved in social cognition, specifically in perpetrating mothers. This neuroimmune pathway may contribute to maladaptive interpretation of child signals during caregiving and represents a potential target for biomarker-informed preventive intervention.

## Introduction

Child maltreatment is a global public health concern with lasting and pervasive effects on children’s physical, psychological, and emotional development (Mehta et al., 2023; Norman et al., 2012; Teicher and Samson, 2016, 2013). To date, mainstream research has predominantly focused on the victims of abuse, aiming to accumulate knowledge that can inform prevention, intervention, and treatment by elucidating the neurobiological consequences of maltreatment (Daníelsdóttir et al., 2024; Goemans et al., 2023). However, to prevent maltreatment at its source and to support struggling caregivers themselves, it is essential to investigate the neurobiological risk factors within the caregivers that lead to abusive behaviors.

Research on caregiver determinants of maltreatment has yielded important insights, particularly from epidemiological studies on socioeconomic status (Esposito et al., 2024; Putnam-Hornstein et al., 2022) and from psychological and psychiatric studies on intergenerational transmission (Assink et al., 2018; Narayan et al., 2021), mental illness (Seeger et al., 2022; Sobral et al., 2025), and neurodevelopmental disorders (Tachibana et al., 2017). While these approaches identify population-level correlates, they offer limited insight into how such factors translate into maltreating behavior. A growing body of research has therefore focused on caregivers’ neurobiology (Carollo et al., 2025; Kaplan et al., 2025; Olsavsky et al., 2021). However, studies have largely examined typical maternal functions such as attachment and empathy (Kim et al., 2016; Orchard et al., 2023) or high-risk groups such as mothers with postpartum depression (Horáková et al., 2022; Morgan et al., 2021; Sobral et al., 2025; Wonch et al., 2016). Because recruiting caregivers who have actually perpetrated maltreatment is particularly challenging, direct neurobiological research in this case group remains scarce.

Against this backdrop, research has begun to conceptualize maltreatment itself as a behavioral dimension and to investigate its neural basis. Rodrigo, León, and colleagues studied mothers identified by Child Protective Services (CPS) for neglect and reported structural and functional abnormalities in brain regions involved in empathy and emotion processing (León et al., 2019; Rodrigo et al., 2020, 2016). These included reduced GMV in the right insular and cingulate cortices using VBM (Rodrigo et al., 2020), decreased connectivity in the right inferior longitudinal fasciculus via DTI (Rodrigo et al., 2016), and blunted amygdala and hippocampal responses to infant emotional stimuli using fMRI (León et al., 2019). These abnormalities were also associated with alexithymia and lower parenting quality (León et al., 2021, 2019; Rodrigo et al., 2016). We recently reported reduced axial diffusivity and fractional anisotropy in the right corticospinal tract in mothers who had perpetrated neglect, physical, and emotional abuse, with these alterations linked to maternal childhood maltreatment and depressive symptoms (Kurata et al., 2024). Together, these findings suggest vulnerabilities across empathy-emotion and motor control systems, but direct evidence in perpetrating caregivers remains limited.

To clarify these neural vulnerabilities and probe their biological mechanisms, we hypothesized that immune-inflammatory dysregulation contributes to structural brain abnormalities. This hypothesis is grounded in evidence linking the psychosocial risks faced by maltreating mothers to inflammation. Mothers’ own childhood maltreatment, commonly reported in this population, can trigger long-lasting peripheral and central inflammatory responses (Andersen, 2022; Beurel et al., 2024; Danese and Baldwin, 2017; Nettis et al., 2020). Chronic psychosocial stressors such as poverty and social isolation also promote pro-inflammatory cytokine production (Iyer et al., 2022; Muscatell et al., 2020), and depression, a risk factor for maltreatment, particularly neglect, is closely linked to inflammatory markers (Mac Giollabhui et al., 2026; Miller and Raison, 2016; Poletti et al., 2024). Chronic inflammation may impair neuroplasticity and brain remodeling through excessive activation of microglia and astrocytes (Calcia et al., 2016; Haroon et al., 2017; Liddelow and Barres, 2017). Consistently, psychiatric MRI studies have linked IL-6 and TNF-α to reduced GMV in prefrontal and temporal cortices and to white matter abnormalities in depression and schizophrenia (Kakeda et al., 2018; Lim et al., 2021; Serpa et al., 2023; Williams et al., 2022). These regions overlap with abnormalities reported in neglectful mothers and core nodes of the empathy-emotion regulation network (Arioli et al., 2021; Rodrigo et al., 2020; Underwood et al., 2021), raising the possibility that immune dysregulation contributes to caregiving-related perpetration through prefrontal-temporal structural alterations.

Although brain structural findings and inflammation-related findings in perpetrating caregivers have begun to emerge, few studies have integrated these strands into a coherent account of parenting difficulties. The present study tested whether structural brain alterations mediate the association between systemic inflammation-related markers and caregiving-related behavioral risk. By integrating structural brain metrics and inflammation-related proteomic markers, we sought to clarify how protective and pro-inflammatory immune processes may shape these vulnerabilities and to identify objective, modifiable targets for future caregiver support and prevention.

## Methods

### Ethics statement

The study protocol was approved by the Ethics Committee of the University of Fukui, Japan (approval numbers 20150109, 20160120, 20220034, and 20220039). All procedures were conducted in accordance with the Declaration of Helsinki and the Ethical Guidelines for Clinical Studies of the Ministry of Health, Labour and Welfare of Japan. All participants provided written informed consent prior to participation.

### Participants

The maltreatment group comprised 16 mothers who met two primary inclusion criteria: 1) a documented history of intervention from CPS or equivalents due to child maltreatment, and 2) confirmation by a consensus of three child psychiatrists (S.K., T.M., and A.T.) that the participant’s history met the study’s specific definition of maltreatment. All mothers in this group had a history of perpetrating at least one form of child maltreatment, including physical abuse, emotional abuse, or neglect. Psychiatric comorbidities included major depressive disorder (*n* = 5), obsessive-compulsive disorder (*n* = 2), bipolar disorder (*n* = 2), post-traumatic stress disorder (*n* = 1), attention-deficit/hyperactivity disorder (ADHD; *n* = 2) and comorbid ADHD and autism spectrum disorder (ASD; *n* = 1). Five mothers in the maltreatment group were taking psychiatric medications at the time of the study. A medication washout was not implemented to avoid potential adverse effects on the children from the mothers’ medication discontinuation. The control group consisted of 145 age-matched mothers recruited from the general population. This group included participants from several of our laboratory’s previous studies. Exclusion criteria for all participants included a history of head trauma with loss of consciousness, significant perinatal or neonatal complications, and any MRI contraindications. A subgroup of participants from both groups (*n* = 53; 11 from the maltreatment group and 42 from the control group) provided blood samples for a separate proteomic analysis. Diagnostic information was available for the index child of each mother in the maltreatment group (*n* = 16). Of these 16 children, 15 had been diagnosed with a neurodevelopmental disorder according to DSM-5 criteria: ADHD (*n* = 2), ASD (*n* = 3), or comorbid ADHD and ASD (*n* = 10). The remaining child had no formal diagnosis. Clinically, it is often challenging to disentangle whether such neurodevelopmental symptoms are temperamental in origin or emerge as sequelae of maltreatment (Capusan et al., 2016; Stern et al., 2018; van der Kolk et al., 2005).

## Measures and Procedures

### Psychological measures

Participants completed several self-report questionnaires, including the Japanese version of the Childhood Trauma Questionnaire (CTQ-J) (Bernstein et al., 1997; Mizuki and Fujiwara, 2021), an Adverse Childhood Experiences (ACEs) scale (Felitti et al., 1998), the Self-Rating Depression Scale (SDS) (Fukuda and Kobayashi, 1973; Zung et al., 1965), the Parenting Scale (PS) (Arnold et al., 1993; Itani, 2010), and the Interpersonal Reactivity Index (IRI) (Davis, 1980; Himichi et al., 2017). Detailed descriptions of these scales and their psychometric properties in this cohort are provided in our previous publications (Kawaguchi et al., 2025; Kurata et al., 2024).

### MRI Acquisition and Pre-processing

All structural brain images were acquired on a 3T PET/MR scanner (Signa PET/MR; GE Healthcare, Milwaukee, WI, USA) with an 8-channel head coil. High-resolution T1-weighted 3D Fast Spoiled Gradient-Recalled Echo (FSPGR) BRAVO images were acquired covering the whole brain. The imaging parameters were as follows: repetition time (TR) = 8.464 ms; echo time (TE) = 3.248 ms; inversion time = 600 ms; flip angle = 11°; voxel size = 1 × 1 × 1 mm³; field of view (FOV) = 256 × 256 mm²; and acquisition matrix = 256 × 256 × 172.

Voxel-based morphometry (VBM) was used to examine voxel-wise regional differences in gray matter volume (GMV). Image pre-processing was conducted using Statistical Parametric Mapping (SPM12; Wellcome Centre for Human Neuroimaging, London, UK) implemented in MATLAB R2022b (MathWorks, Natick, MA, USA). T1-weighted images were segmented into gray matter, white matter, and cerebrospinal fluid probability maps. The resulting gray matter images were then normalized to the Montreal Neurological Institute (MNI) standard space using the DARTEL (Diffeomorphic Anatomical Registration Through Exponentiated Lie Algebra) algorithm. Modulation preserved local tissue volumes following spatial normalization. Finally, the normalized gray matter images were smoothed with an 8-mm full width at half maximum (FWHM) Gaussian kernel.

In addition to the T1-weighted images, diffusion tensor imaging (DTI) data were acquired during the same scanning session for a subgroup of participants. The primary analysis of these DTI data, including group differences in white matter microstructure, has been reported previously (Kurata et al., 2024). The present study used these previously identified white matter findings as the basis for a secondary exploratory cross-modal analysis.

### Plasma Collection and Proteomic Pre-processing

For a subgroup of participants (*n* = 53), whole blood samples were collected in EDTA-K2 tubes (Venoject II, VP-DK050K, Terumo Corporation, Tokyo, Japan). Plasma was immediately isolated by centrifugation, aliquoted, and stored at −80°C until analysis. The concentrations of 92 inflammation-related proteins were measured using the Olink Target 96 Inflammation panel (Olink Proteomics, Uppsala, Sweden), which is based on Proximity Extension Assay (PEA) technology (Assarsson et al., 2014).

Data quality control was performed according to Olink’s standard protocol. One sample was identified as a technical outlier due to significant deviations in its internal controls and was subsequently excluded (Supplementary Figure S1; final *n* = 52). Twelve proteins were omitted because more than 50% of their measurements fell below the lower limit of detection (LLOD). For the remaining 80 proteins, any values below the LLOD were imputed using the GSimp method (Wei et al., 2018). Three proteins (ST1A1, AXIN1, and CXCL5) with non-normal distributions were transformed using a Rank-based Inverse Normal Transformation (RINT). The final protein expression levels are presented in log_2_-transformed Normalized Protein eXpression (NPX) units.

### Statistical Analysis

All statistical analyses were performed using R version 4.4.2 (R Core Team, 2025). To account for multiple comparisons, we used methods to control the false discovery rate (*FDR*) at a 5% threshold, as detailed for each analysis below.

### Group Comparisons

To assess group differences in brain structure, VBM analyses of GMV were performed using a voxel-wise multiple regression model within the general linear model (GLM) framework in SPM12, comparing the maltreatment group (*n* = 16) to the control group (*n* = 145). Age was included as a covariate of no interest, and regional GMV was normalized using proportional scaling relative to total intracranial volume (TIV) (Ashburner and Friston, 2000). The statistical threshold was set at *p* < 0.05, *FDR*-corrected at the cluster level. Peak-voxel statistics within significant clusters are reported for reference. Anatomical labels for significant clusters were assigned using the Neuromorphometrics atlas (RRID:SCR_005656; Neuromorphometrics, Inc.).

To identify proteins with differential expression levels between groups, a robust linear regression model was fitted for each of the 80 proteins, with group as the predictor of interest and age as a covariate. The resulting *p*-values, derived from the model’s *t*-statistics, were then converted to *q*-values using Storey’s method (Storey, 2002) to control the *FDR*. A *q*-value < 0.05 was considered statistically significant.

### Correlation Analyses

To examine relationships between the gray matter alterations and other multimodal data, we extracted mean β-values (representing the group difference effect size) from the significant VBM cluster and used these in subsequent correlation analyses. First, to relate gray matter findings to our previously published white matter alterations (Kurata et al., 2024), we performed a cross-modal analysis in the same subgroup used in our previous DTI study (*n* = 51; 11 maltreatment and 40 control), which partially overlaps with the proteomic subgroup. VBM-derived β-values were tested for associations with mean FA and AD values extracted from the previously identified right corticospinal tract (R.CST) cluster, as these diffusion metrics had shown significant between-group differences in the prior report. The *p*-values for these two tests were corrected for multiple comparisons using the Benjamini-Hochberg (BH) method to control the *FDR*. Second, we examined the associations between brain structure and behavioral/biological measures. The VBM β-values were correlated with psychological measures. We also conducted a targeted correlation between the VBM β-values and the expression levels of proteins that had been found to be significantly different between the groups in our primary analysis. Prior to this analysis, the protein expression levels were corrected for the effect of age, and these age-adjusted values were used in the correlations. The *p*-values were corrected using Storey’s method for *FDR* control (*q* < 0.05).

### Mediation analysis

We performed mediation analyses to test whether peripheral immune marker profiles were associated with caregiving-related perpetration through structural brain variation. The mediator was defined as the mean GMV extracted from the cluster showing a significant between-group difference in the whole-brain VBM analysis. The independent variables were two composite immune indices derived from distinct sets of proteins that showed both significant group differences and significant associations with the extracted GMV measure. Our analytic strategy followed a sequential, hypothesis-driven design. Building on the directional framework outlined in the Introduction—namely, that immune dysregulation is linked to perpetration through structural brain alterations—we conducted three component analyses to characterize the constituent bivariate relationships: (i) group differences in GMV (group → GMV), (ii) group differences in proteins (group → proteins), and (iii) protein–GMV associations (proteins → GMV). The directionality of each path observed in these component analyses was consistent with our a priori hypotheses (see Results). Mediation analyses were therefore conducted as a hypothesis-confirming integrative test of the indirect pathway, rather than as an exploratory analysis. Before mediation testing, logistic regression models were fitted with perpetration status as the binary outcome to confirm independent associations of each immune composite, both separately and simultaneously. Separate mediation models were then estimated for each immune composite. In each model, path *a* represented the association between the immune composite and GMV, and path *b* represented the association between GMV and perpetration status. Given the directional hypotheses derived from prior literature and supported by the preceding component analyses, indirect effects were evaluated using one-tailed tests with 90% confidence intervals, in line with recommendations for hypothesis-driven analyses with pre-specified directionality (Lakens et al., 2018). Indirect effects were quantified as the product of coefficients (*a* × *b*) and were considered significant when the confidence interval excluded zero. Models were estimated within a structural equation modeling framework using the R package *lavaan* (Rosseel, 2012) with the WLSMV estimator, which accommodates binary outcomes.

## Results

### Demographic characteristics

Demographic characteristics of the maltreatment and control groups are summarized in Table 1. There were no significant group differences in age or number of children (Welch’s *t*-test: age, *t*(17) = −1.20, *p* = 0.25; number of children, *t*(16) = −1.20, *p* = 0.24). In contrast, cohabitation status differed significantly between groups (Fisher’s exact test, *p* < 0.001), with substantially higher odds of being single in the maltreatment group (OR = 15.4, 95% CI [4.0, 64.7]).

**Table 1.**
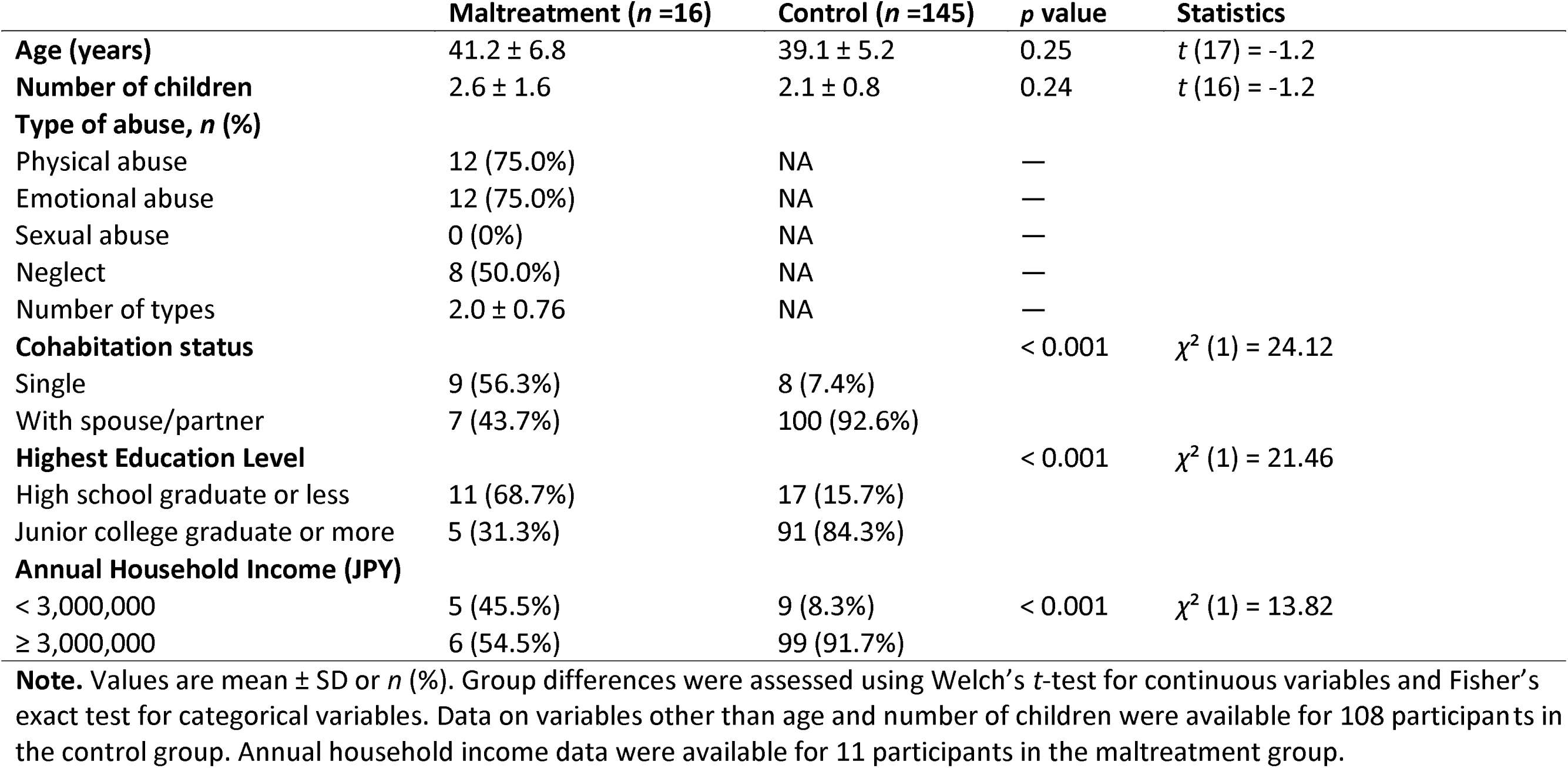
Demographics of the participants.

Highest education level also differed significantly between groups (Fisher’s exact test, *p* < 0.001), with higher odds of having a high school education or less in the maltreatment group (OR = 11.4, 95% CI [3.2, 47.7]). Annual household income was summarized descriptively because income data were available for only 11 participants in the maltreatment group.

### Voxel-based morphometry (VBM)

The whole-brain VBM analysis revealed a single significant cluster where the maltreatment group exhibited reduced GMV compared to the control group (cluster-level *q* = 0.020, k = 506 voxels; peak MNI: x = 64, y = -38, z = -18; *T* = 3.96, peak-level *q* = 0.113), corresponding to the right middle and inferior temporal gyri (hereafter, R.MTG/ITG; Figure 1A). No brain regions showed significantly greater GMV in the maltreatment group. The effect size for this group difference was large (Cohen’s *d* = -1.01). A post-hoc power analysis confirmed adequate statistical power (>0.97) to detect an effect of this magnitude despite the imbalanced sample sizes (16 vs 145).

**Figure 1.**
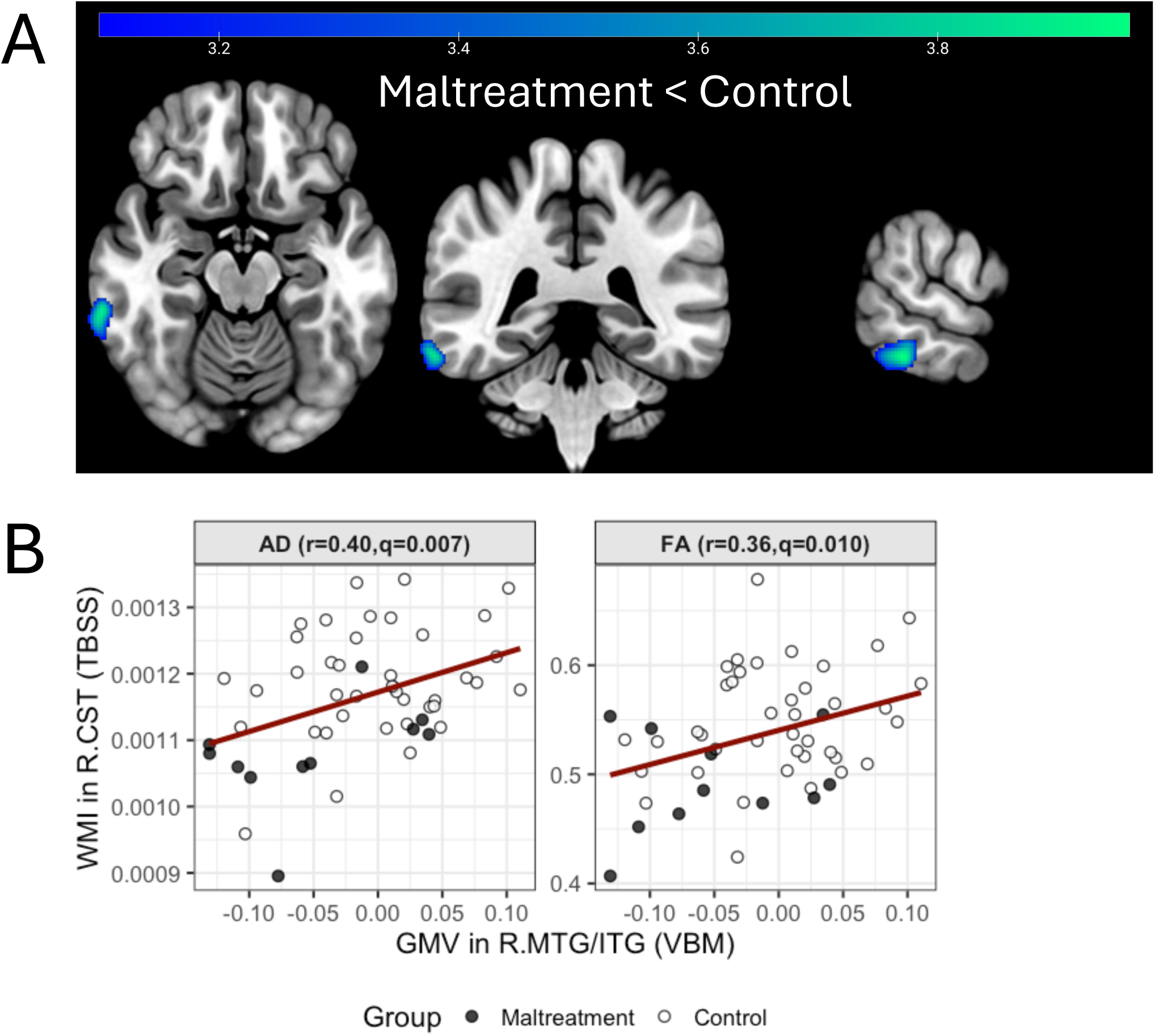
Reduced temporal GMV in maltreating mothers and its association with corticospinal tract microstructure. **(A)** A VBM analysis shows a significant cluster of reduced GMV in the right middle/inferior temporal gyrus (R.MTG/ITG) in the maltreatment group versus controls (cluster-level *FDR*-corrected *q* = 0.020). **(B)** In a subsample with multimodal data (*n* = 50), GMV from the R.MTG/ITG cluster was significantly correlated with diffusion metrics from the right corticospinal tract (R.CST), previously identified in Kurata et al. (2024). Lower GMV was associated with lower axial diffusivity (AD) and fractional anisotropy (FA).

### Association with White Matter Integrity (WMI)

We next examined associations between R.MTG/ITG GMV and the two conventional DTI metrics (AD and FA) previously reported in the R.CST cluster (Kurata et al., 2024). After correcting for multiple comparisons, GMV in the R.MTG/ITG showed significant positive correlations with both AD (*r* = 0.40, *q* = 0.007) and FA (*r* = 0.36, *q* = 0.010) in the R.CST (Figure 1B).

### Differential Protein Expression

After controlling for the *FDR*, 16 proteins showed significantly different expression levels between the maltreating and control mothers (all *q* < 0.05) (Figure 2, Supplementary Table S1). The 16 proteins were almost evenly split between up- and down- regulation in the maltreatment group. Specifically, the maltreatment group exhibited significantly higher levels of seven proteins (up-regulated), encompassing pro-inflammatory cytokines (IL-17C, IL-17A, CCL20) and immune-related markers (CD5, IL-15RA, SLAMF1, IL-10RB). Conversely, the maltreatment group showed significantly lower levels of nine down-regulated proteins, encompassing chemokines (CXCL1, CXCL6, MCP-2, MCP-4), regulatory and metabolic proteins (SIRT2, STAMBP, 4E-BP1), and a TNF-superfamily member (TWEAK), as well as a neurotrophin (NT-3).

**Figure 2.**
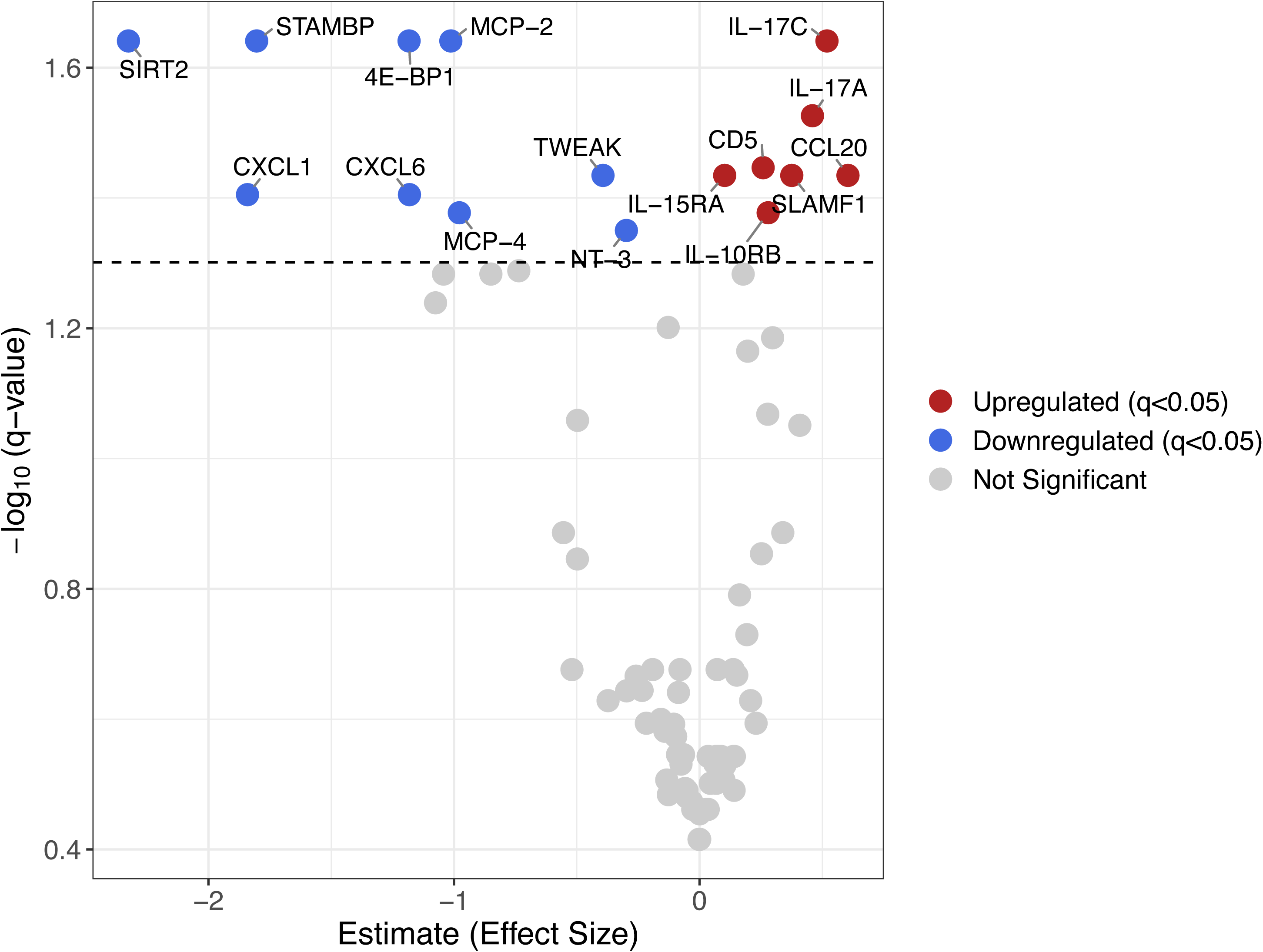
Altered plasma inflammatory protein profile in maltreating mothers. Volcano plot of 80 inflammation-related proteins comparing maltreating mothers to controls. Sixteen proteins were significantly differentially expressed (*q* < 0.05, *FDR*-corrected), with seven upregulated (red) and nine downregulated (blue) in the maltreatment group. The x-axis represents the effect size, and the y-axis represents the statistical significance (-log_10_ *q*-value).

### Association between GMV in the R.MTG/ITG and Inflammatory Proteins

After *FDR* correction, nine of the 16 differentially expressed proteins remained significantly correlated with R.MTG/ITG GMV (Figure 3, Supplementary Table S2). These significant correlations included both positive and negative associations. Specifically, reduced GMV was correlated with higher levels of CCL20 (*r* = -0.43) and IL-17C (*r* = -0.37). Conversely, relatively preserved GMV was correlated with higher levels of CXCL1 (*r* = 0.41), CXCL6 (*r* = 0.37), SIRT2 (*r* = 0.36), STAMBP (*r* = 0.35), MCP-2 (*r* = 0.32), MCP-4 (*r* = 0.30), and 4E-BP1 (*r* = 0.30). A subsequent exploratory analysis found no significant correlations between the two DTI metrics (from the R.CST) and the 16 proteins after correcting for multiple comparisons.

**Figure 3.**
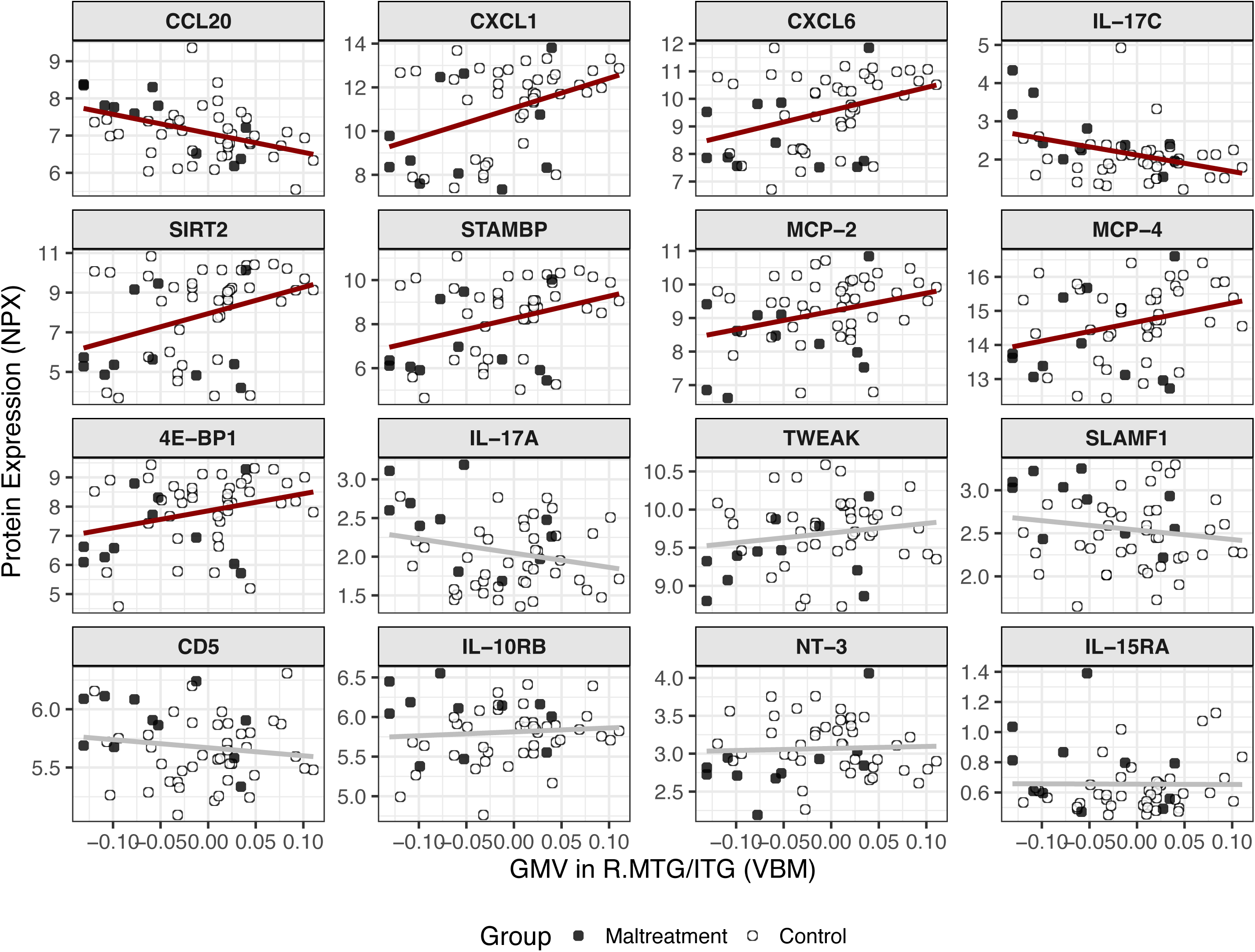
Temporal GMV is correlated with peripheral inflammatory markers. Scatter plots showing the correlation between GMV in the right temporal cluster (R.MTG/ITG) and the 16 proteins that were differentially expressed between groups. After *FDR* correction, nine proteins showed a significant association with GMV (indicated by red lines; *q* < 0.05).

### Immune system–level associations with caregiving-related perpetration and mediation through R.MTG/ITG GMV

Logistic regression analyses demonstrated that the upregulated immune set (CCL20 and IL-17C) was significantly associated with caregiving-related perpetration (OR = 2.15, 95% CI [1.13–4.75], *p* = 0.028), whereas the downregulated immune set (CXCL1, CXCL6, MCP-2, MCP-4, SIRT2, STAMBP, and 4E-BP1) showed a significant inverse association (OR = 0.41, 95% CI [0.17–0.86], *p* = 0.027). When both sets were simultaneously included in the model, these opposing effects remained significant, indicating independent risk (upregulated) and buffering (downregulated) pathways. Because these logistic regression results, together with the preceding bivariate correlations and VBM group comparisons, confirmed the hypothesized directionality for all component paths, indirect effects in the subsequent mediation analyses were evaluated using one-tailed tests, as prespecified in the Methods.

Mediation analyses were then performed to test whether R.MTG/ITG GMV mediated these immune–perpetration associations (Figure 4). For the upregulated set, a higher composite score was associated with lower R.MTG/ITG GMV (path *a*: β = −0.46), and lower GMV was associated with perpetration status (path *b*: β = −0.33). The indirect effect was significant (*a* × *b* = 0.167, *p* = 0.049, 90% CI [0.001, 0.332]). For the downregulated set, a higher composite score was associated with greater R.MTG/ITG GMV (path *a*: β = 0.38), while GMV was again negatively associated with perpetration status (path *b*: β = −0.36). The indirect effect was also significant (*a* × *b* = −0.152, *p* = 0.038, 90% CI [−0.292, −0.011]).

**Figure 4.**
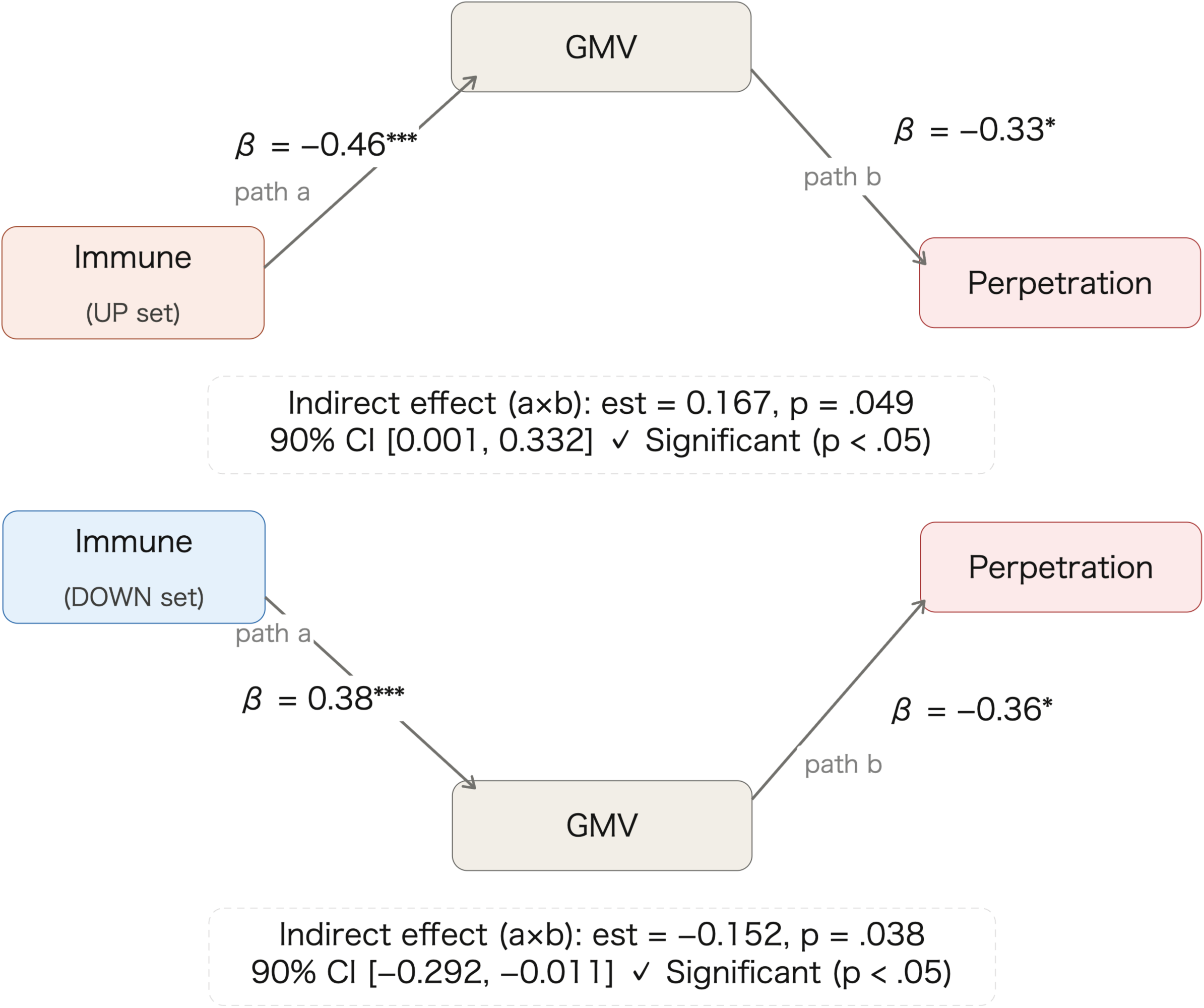
Indirect associations of peripheral immune marker sets with perpetration status via right temporal gray matter volume. Mediation models testing whether GMV in the R.MTG/ITG mediates the associations between peripheral immune marker sets and caregiving-related perpetration. The upper panel shows the model for the upregulated immune set (Up set: CCL20 and IL-17C), and the lower panel shows the model for the downregulated immune set (Down set: CXCL1, CXCL6, SIRT2, STAMBP, MCP-2, MCP-4, and 4E-BP1). Path *a* represents the association between each immune composite and R.MTG/ITG GMV; path *b* represents the association between GMV and perpetration status. Indirect effects (*a* × *b*) were evaluated using one-tailed *p* values and 90% confidence intervals. Standardized path coefficients (β) are shown. \**p* < .05, \*\*\**p* < .001. GMV, gray matter volume; R.MTG/ITG, right middle/inferior temporal gyrus.

## Discussion

The present study examined whether peripheral immune imbalance in mothers who perpetrated child maltreatment is associated with structural brain alterations, and whether such alterations mediate the link to perpetration. Integrating VBM-derived structural brain measures, inflammation-related proteomic markers, and psychological indices, we observed reduced GMV in the R.MTG/ITG in the maltreatment group. Several inflammation-related proteins that differed between groups were also associated with GMV in this region. In addition, immune markers showing opposite directional profiles showed independent associations with perpetration. Together, these findings provide preliminary support for an immune–neural–behavioral framework of maltreatment perpetration, suggesting that this behavior cannot be reduced to a single psychosocial factor.

One interpretation of the immune profile in the maltreatment group is that it reflects biological embedding of prolonged psychosocial stress. Mothers in this group have previously shown elevated adverse childhood experiences (ACE and CTQ) and depressive symptoms (SDS) (Kawaguchi et al., 2025). In the subset completing the Parenting Stress Index (PSI) (Abidin, Richard R., 1995; Narama, M. et al., 1999), caregiving stress was also higher in the maltreatment group (261.3 ± 35.2 vs. 169.9 ± 33.7; Welch’s t-test: t(4) = −4.84, p = 0.008, Cohen’s d = 2.69), exceeding the clinically significant threshold despite the small subgroup. This is consistent with prior reports of elevated caregiving stress among parents at risk for or engaged in maltreating behaviors (Crouch and Behl, 2001; Rodriguez and Green, 1997). Chronic stress is linked to altered peripheral immune signaling (Iyer et al., 2022; Muscatell et al., 2020). Importantly, the immune alterations were not limited to inflammatory elevations: IL-17C and CCL20 co-occurred with reductions in proteins involved in immune regulation, intracellular signaling, or immune cell trafficking, including CXCL1, CXCL6, MCP-2, MCP-4, SIRT2, STAMBP, and 4E-BP1 (Bednash et al., 2021; He et al., 2020; Huppert et al., 2010; Palomino and Marti, 2015; Pearl et al., 2020; Reboldi et al., 2009; Sasso et al., 2014). Thus, disrupted balance between immune drive and immune control, rather than simply high or low inflammation, may be relevant to perpetration.

The reduction in R.MTG/ITG GMV may provide insight into neural processes relevant to caregiving behavior. Because the significant cluster crossed anatomical boundaries, we interpret the finding at the level of right lateral temporal association cortex rather than either subregion alone. Within this territory, the MTG is implicated in controlled semantic retrieval and event-related semantic processing, whereas the inferior temporal region is linked to higher-order visual representations of complex objects and faces (Conway, 2018; Davey et al., 2016; Ishai et al., 1999; Whitney et al., 2011). The present result may therefore involve an interface between higher-order perceptual representation and contextual/social meaning construction. In caregiving, parents must rapidly interpret ambiguous child cues such as crying, facial expression, gaze, defiance, or withdrawal. Under stress, limited attentional and regulatory resources may increase reliance on fragmentary or negatively weighted information, making a child’s behavior more likely to be experienced as provocation than as distress or need. Reduced GMV in this cluster may therefore reflect vulnerability in lateral temporal processes relevant to interpretation of child-related cues, although this remains inferential because these operations were not directly measured. Prior work shows that abuse-risk or stressed parents more often interpret ambiguous child behavior negatively, and such attributions are associated with harsh or dysfunctional parenting (Beckerman et al., 2018; Dadds et al., 2003). This interpretation is also consistent with the Social Information Processing model, in which biases during perception and interpretation of child behavior are associated with abusive parenting (Camilo et al., 2020; Milner, 2003, 1993). The present findings may extend this framework by linking these psychological biases to immune and neurobiological substrates.

The present results may also complement previous neuroimaging studies emphasizing emotional processing alterations in maltreating caregivers. Rodrigo and colleagues reported reduced GMV in the insula, cingulate cortex, and inferior frontal gyrus among neglectful caregivers, suggesting alterations in neural systems involved in salience processing, interoceptive awareness, and emotion regulation (Rodrigo et al., 2020). In contrast, the present finding in right lateral temporal association cortex may point more strongly to a cognitive–social interpretative component involved in construing the meaning of child signals.

An additional issue concerns the association between the VBM finding and the white matter indices identified in our previous TBSS analysis, particularly in the R.CST (Kurata et al., 2024). Because VBM-derived GMV and TBSS-derived diffusion metrics reflect distinct structural properties of gray and white matter, respectively, we do not interpret this association as evidence of a direct anatomical or mechanistic linkage. Rather, such covariation across individuals may be informative at the systems level, as both modalities may be influenced by shared developmental, stress-related, or neuroimmune factors. In this sense, the observed association between the R.MTG/ITG cluster and the R.CST should be regarded as an exploratory cross-modal covariance finding. Immune markers were not significantly associated with R.CST measures, which may suggest that the peripheral immune profile identified here is more closely related to gray matter variation in temporal associative cortex than to this white matter pathway. Because no complementary multimodal analyses, such as functional connectivity, tractography, or integrated multivariate modeling, were performed in the present study, these findings cannot yet be interpreted in an integrated network-level manner. This interpretation should therefore be considered tentative, pending future studies that can determine whether these cross-modal associations reflect shared upstream factors or coordinated network-level vulnerability.

The mechanisms linking peripheral immune imbalance and structural brain variation remain speculative but may involve neuroimmune communication pathways. Peripheral inflammatory signals can affect central nervous system function through humoral routes via the blood–brain barrier and circumventricular organs, and may also be conveyed through vagal afferent pathways projecting to the brainstem and limbic structures (Dantzer et al., 2008; Hosoi et al., 2000; Pavlov and Tracey, 2017; Quan and Banks, 2007). These pathways are not mutually exclusive; under chronic stress, repeated peripheral immune input may alter microglial reactivity and promote sustained neuroinflammatory states, potentially leading to excessive synaptic pruning, reduced neuropil density, and lower cortical GMV (C.Paolicelli et al., 2011; Hong et al., 2016; Schafer et al., 2012). The elevated IL-17C and CCL20 observed in the present study are compatible with enhanced Th17-related inflammatory signaling (Bettelli et al., 2008; Korn et al., 2009; McGeachy et al., 2019; Reboldi et al., 2009) and immune cell trafficking, whereas co-occurring reductions in chemokines and intracellular regulatory proteins suggest weakened homeostatic control within immune networks. Thus, a state in which inflammatory drive and regulatory decline coexist may converge on structural vulnerability in temporal association cortex. Consistent with this view, inflammatory pathways have been reported to show regionally selective associations with brain structure, including in temporal cortical regions (Bao et al., 2024; Gu et al., 2017; Jacomb et al., 2018; Williams et al., 2022). These observations support the broader notion that peripheral immune imbalance may be relevant to regionally selective brain vulnerability.

Several limitations should be acknowledged. First, the cross-sectional design precludes conclusions regarding temporal ordering or causality among immune imbalance, brain structural alterations, and caregiving-related behaviors. It remains possible that chronic psychosocial stress is associated with both immune and neural alterations in parallel, rather than through a sequential pathway. Accordingly, the mediation models presented here should be interpreted as hypothesis-generating statistical models that are consistent with the observed associations, rather than as evidence of causal mechanisms. Second, the sample size of the maltreatment group was relatively small, increasing uncertainty in parameter estimates and raising concerns regarding effect-size inflation and replicability. Third, although age was included as a covariate, several factors that may affect immune markers—including socioeconomic status, allergic conditions, medication use, substance misuse, and recent infection history—were not fully controlled in the present analyses. Fourth, the immune markers examined here were derived from peripheral blood and do not directly index neuroinflammatory processes within the central nervous system. Future work should include independent replication, longitudinal designs with repeated measures, more refined control of confounding variables, and, where feasible, multilevel verification incorporating central nervous system indices.

Despite these limitations, the present study provides preliminary evidence consistent with a framework in which peripheral immune imbalance—characterized by increased inflammatory drive alongside weakened regulatory processes—is associated with structural variation in the temporal association cortex. Such variation may be relevant to how caregivers interpret socially ambiguous child signals and may thereby contribute to vulnerability for maladaptive caregiving. Importantly, the neurobiological characteristics identified in this study suggest that maltreating behavior should not necessarily be viewed solely as a fixed trait of caregivers. Rather, such behavior may emerge from a convergence of psychosocial factors—such as the caregiver’s own childhood adversity and current stress—converge with biological factors, including the atypical brain structure and protein expression profiles identified here, against the broader context of socially and culturally imposed responsibilities and the high physical and temporal demands of child-rearing. Some caregivers who engage in abusive behaviors report difficulty controlling their reactions and experience guilt while simultaneously expressing a desire to care for and protect their children (Azar et al., 2008; Crouch et al., 2017, 2008; Dym Bartlett and Easterbrooks, 2015) Recognizing this complexity may help shift the perspective from viewing abusive caregivers solely as targets of blame toward understanding them as individuals who may also require support and intervention. This perspective does not minimize the serious consequences of maltreatment for children; rather, it emphasizes that effective prevention may require identifying modifiable vulnerabilities in caregivers alongside psychosocial risk conditions. From a translational perspective, if future longitudinal and intervention studies support the present framework, integrating immune, neural, and psychosocial indicators may contribute to more individualized prevention strategies. Such approaches may help refine risk stratification and guide interventions that combine psychosocial care with attention to physical health and lifestyle factors. More broadly, the present findings raise the possibility that approaches aimed at reducing chronic stress, restoring physiological regulation, and improving social-cognitive interpretation of child signals—always prioritizing the safety of both children and caregivers—may contribute to building prevention and support systems that foster the healthy development of children.

## Data Availability

All data produced in the present study are available upon reasonable request to the authors

## Acknowledgments

We thank the Research Center for Child Mental Development and the Department of Child and Adolescent Psychological Medicine at the University of Fukui Hospital for their generous assistance with participant recruitment and data collection.

## Author contributions

SK, SN (2^nd^), and AT conceived and designed the project. SK, SN (2^nd^), NYSK, AY, RK (5^th^), RK (6^th^), SN (7^th^), TM, TXF, YF, HO, and AT collected the data. SK, SN (2^nd^), NYSK, and TXF analyzed and interpreted the data. SK and SN (2^nd^) drafted the manuscript with critical revisions and supervision from TM, TXF, YF, HO, and AT. All authors have approved the final manuscript.

## Data availability statement

The datasets generated and/or analyzed during the current study are not publicly available due to the risk of participant identification and the need to protect participant anonymity. However, the data are available from the corresponding authors on reasonable request.

## Declaration of generative AI use

During the preparation of this work, the authors used ChatGPT (OpenAI) to support English language editing and grammar checking. After using this tool, the authors reviewed and edited the content as needed and take full responsibility for the content of the publication.

## References

Arnold, D.S., O’Leary, S.G., Wolff, L.S., Acker, M.M., 1993. The Parenting Scale: A measure of dysfunctional parenting in discipline situations. Psychol. Assess. 5, 137–144. 10.1037/1040-3590.5.2.137

Ashburner, J., Friston, K.J., 2000. Voxel-based morphometry--the methods. NeuroImage 11, 805–821. 10.1006/nimg.2000.0582

Assarsson, E., Lundberg, M., Holmquist, G., Björkesten, J., Thorsen, S.B., Ekman, D., Eriksson, A., Rennel Dickens, E., Ohlsson, S., Edfeldt, G., Andersson, A.-C., Lindstedt, P., Stenvang, J., Gullberg, M., Fredriksson, S., 2014. Homogenous 96-plex PEA immunoassay exhibiting high sensitivity, specificity, and excellent scalability. PloS One 9, e95192. 10.1371/journal.pone.0095192

Bao, Y., Chen, X., Li, Y., Yuan, S., Han, L., Deng, X., Ran, J., 2024. Chronic Low-Grade Inflammation and Brain Structure in the Middle-Aged and Elderly Adults. Nutrients 16, 2313. 10.3390/nu16142313

Bernstein, D.P., Ahluvalia, T., Pogge, D., Handelsman, L., 1997. Validity of the Childhood Trauma Questionnaire in an adolescent psychiatric population. J. Am. Acad. Child Adolesc. Psychiatry 36, 340–348. 10.1097/00004583-199703000-00012

Bettelli, E., Korn, T., Oukka, M., Kuchroo, V.K., 2008. Induction and effector functions of T(H)17 cells. Nature 453, 1051–1057. 10.1038/nature07036

Capusan, A.J., Kuja-Halkola, R., Bendtsen, P., Viding, E., McCrory, E., Marteinsdottir, I., Larsson, H., 2016. Childhood maltreatment and attention deficit hyperactivity disorder symptoms in adults: a large twin study. Psychol. Med. 46, 2637–2646. 10.1017/S0033291716001021

C.Paolicelli, R., Giulia Bolasco, Francesca Pagani, Laura Maggi, Maria Scianni, Patrizia Panzanelli, Maurizio Giustetto, Tiago Alves Ferreira, Eva Guiducci, Laura Dumas, Davide Ragozzino, Cornelius T. Gross, 2011. Synaptic Pruning by Microglia Is Necessary for Normal Brain Development. Science 333, pp.1456–1458. DOI:%2010.1126/science.1202529

Daníelsdóttir, H.B., Aspelund, T., Shen, Q., Halldorsdottir, T., Jakobsdóttir, J., Song, H., Lu, D., Kuja-Halkola, R., Larsson, H., Fall, K., Magnusson, P.K.E., Fang, F., Bergstedt, J., Valdimarsdóttir, U.A., 2024. Adverse Childhood Experiences and Adult Mental Health Outcomes. JAMA Psychiatry 81, 586–594. 10.1001/jamapsychiatry.2024.0039

Dantzer, R., O’Connor, J.C., Freund, G.G., Johnson, R.W., Kelley, K.W., 2008. From inflammation to sickness and depression: when the immune system subjugates the brain. Nat. Rev. Neurosci. 9, 46–56. 10.1038/nrn2297

Davis, M., 1980. A multidimensional approach to individual differences in empathy.

Felitti, V.J., Anda, R.F., Nordenberg, D., Williamson, D.F., Spitz, A.M., Edwards, V., Koss, M.P., Marks, J.S., 1998. Relationship of childhood abuse and household dysfunction to many of the leading causes of death in adults. The Adverse Childhood Experiences (ACE) Study. Am. J. Prev. Med. 14, 245–258. 10.1016/s0749-3797(98)00017-8

Fukuda, K., Kobayashi, S., 1973. [A study on a self-rating depression scale (author’s transl)]. Seishin Shinkeigaku Zasshi 75, 673–679.

Goemans, A., Viding, E., McCrory, E., 2023. Child Maltreatment, Peer Victimization, and Mental Health: Neurocognitive Perspectives on the Cycle of Victimization. Trauma Violence Abuse 24, 530–548. 10.1177/15248380211036393

Gu, Y., Vorburger, R., Scarmeas, N., Luchsinger, J.A., Manly, J.J., Schupf, N., Mayeux, R., Brickman, A.M., 2017. Circulating inflammatory biomarkers in relation to brain structural measurements in a non-demented elderly population. Brain. Behav. Immun. 65, 150–160. 10.1016/j.bbi.2017.04.022

Himichi, T., Osanai, H., Goto, T., Fujita, H., Kawamura, Y., Davis, M.H., Nomura, M., 2017. [Development of a Japanese version of the Interpersonal Reactivity Index]. Shinrigaku Kenkyu 88, 61–71. 10.4992/jjpsy.88.15218

Hong, S., Beja-Glasser, V.F., Nfonoyim, B.M., Frouin, A., Li, S., Ramakrishnan, S., Merry, K.M., Shi, Q., Rosenthal, A., Barres, B.A., Lemere, C.A., Selkoe, D.J., Stevens, B., 2016. Complement and microglia mediate early synapse loss in Alzheimer mouse models. Science 352, 712–716. 10.1126/science.aad8373

Hosoi, T., Okuma, Y., Nomura, Y., 2000. Electrical stimulation of afferent vagus nerve induces IL-1beta expression in the brain and activates HPA axis. Am. J. Physiol. Regul. Integr. Comp. Physiol. 279, R141–147. 10.1152/ajpregu.2000.279.1.R141

Itani, T., 2010. [The Japanese version of the Parenting Scale: factor structure and psychometric properties]. Shinrigaku Kenkyu 81, 446–452. 10.4992/jjpsy.81.446

Jacomb, I., Stanton, C., Vasudevan, R., Powell, H., O’Donnell, M., Lenroot, R., Bruggemann, J., Balzan, R., Galletly, C., Liu, D., Weickert, C.S., Weickert, T.W., 2018. C-Reactive Protein: Higher During Acute Psychotic Episodes and Related to Cortical Thickness in Schizophrenia and Healthy Controls. Front. Immunol. 9. 10.3389/fimmu.2018.02230

Kawaguchi, Y., Kurata, S., Kawata, N.Y.S., Yao, A., Nishitani, S., Fujisawa, T.X., Tomoda, A., 2025. Effects of childhood maltreatment on mothers’ empathy and parenting styles in intergenerational transmission. Sci. Rep. 15, 7787. 10.1038/s41598-025-92804-0

Korn, T., Bettelli, E., Oukka, M., Kuchroo, V.K., 2009. IL-17 and Th17 Cells. Annu. Rev. Immunol. 27, 485–517. 10.1146/annurev.immunol.021908.132710

Kurata, S., Nishitani, S., Kawata, N.Y.S., Yao, A., Fujisawa, T.X., Okazawa, H., Tomoda, A., 2024. Diffusion tensor imaging of white-matter structural features of maltreating mothers and their associations with intergenerational chain of childhood abuse. Sci. Rep. 14, 5671. 10.1038/s41598-024-53666-0

Lakens, D., Scheel, A.M., Isager, P.M., 2018. Equivalence Testing for Psychological Research: A Tutorial. Adv. Methods Pract. Psychol. Sci. 1, 259–269. 10.1177/2515245918770963

McGeachy, M.J., Cua, D.J., Gaffen, S.L., 2019. The IL-17 family of cytokines in health and disease. Immunity 50, 892–906. 10.1016/j.immuni.2019.03.021

Mehta, D., Kelly, A.B., Laurens, K.R., Haslam, D., Williams, K.E., Walsh, K., Baker, P.R.A., Carter, H.E., Khawaja, N.G., Zelenko, O., Mathews, B., 2023. Child Maltreatment and Long-Term Physical and Mental Health Outcomes: An Exploration of Biopsychosocial Determinants and Implications for Prevention. Child Psychiatry Hum. Dev. 54, 421–435. 10.1007/s10578-021-01258-8

Mizuki, R., Fujiwara, T., 2021. Validation of the Japanese version of the Childhood Trauma Questionnaire-Short Form (CTQ-J). Psychol. Trauma Theory Res. Pract. Policy 13, 537–544. 10.1037/tra0000972

Norman R.E., Byambaa M., De R., Butchart A., Scott J., Vos T., 2012. The Long-Term Health Consequences of Child Physical Abuse, Emotional Abuse, and Neglect: A Systematic Review and Meta-Analysis. PLOS Med. 9, e1001349. 10.1371/journal.pmed.1001349

Pavlov, V.A., Tracey, K.J., 2017. Neural regulation of immunity: molecular mechanisms and clinical translation. Nat. Neurosci. 20, 156–166. 10.1038/nn.4477

Quan, N., Banks, W.A., 2007. Brain-immune communication pathways. Brain. Behav. Immun. 21, 727–735. 10.1016/j.bbi.2007.05.005

R Core Team, 2025. R A Language and Environment for Statistical Computing. R Foundation for Statistical Computing [WWW Document]. URL https://www.scirp.org/reference/referencespapers?referenceid=3967248 (accessed 1.22.26).

Reboldi, A., Coisne, C., Baumjohann, D., Benvenuto, F., Bottinelli, D., Lira, S., Uccelli, A., Lanzavecchia, A., Engelhardt, B., Sallusto, F., 2009. C-C chemokine receptor 6-regulated entry of TH-17 cells into the CNS through the choroid plexus is required for the initiation of EAE. Nat. Immunol. 10, 514–523. 10.1038/ni.1716

Rodrigo, M.J., León, I., García-Pentón, L., Hernández-Cabrera, J.A., Quiñones, I., 2020. Neglectful maternal caregiving involves altered brain volume in empathy-related areas. Dev. Psychopathol. 32, 1534–1543. 10.1017/S0954579419001469

Rosseel, Y., 2012. lavaan: An R Package for Structural Equation Modeling. J. Stat. Softw. 48, 1–36. 10.18637/jss.v048.i02

Schafer, D.P., Lehrman, E.K., Kautzman, A.G., Koyama, R., Mardinly, A.R., Yamasaki, R., Ransohoff, R.M., Greenberg, M.E., Barres, B.A., Stevens, B., 2012. Microglia Sculpt Postnatal Neural Circuits in an Activity and Complement-Dependent Manner. Neuron 74, 691–705. 10.1016/j.neuron.2012.03.026

Stern, A., Agnew-Blais, J., Danese, A., Fisher, H.L., Jaffee, S.R., Matthews, T., Polanczyk, G.V., Arseneault, L., 2018. Associations between abuse/neglect and ADHD from childhood to young adulthood: A prospective nationally-representative twin study. Child Abuse Negl. 81, 274–285. 10.1016/j.chiabu.2018.04.025

Storey, J.D., 2002. A Direct Approach to False Discovery Rates. J. R. Stat. Soc. Ser. B Stat. Methodol. 64, 479–498. 10.1111/1467-9868.00346

Teicher, M.H., Samson, J.A., 2016. Annual Research Review: Enduring neurobiological effects of childhood abuse and neglect. J. Child Psychol. Psychiatry 57, 241–266. 10.1111/jcpp.12507

Teicher, M.H., Samson, J.A., 2013. Childhood maltreatment and psychopathology: A case for ecophenotypic variants as clinically and neurobiologically distinct subtypes. Am. J. Psychiatry 170, 1114–1133. 10.1176/appi.ajp.2013.12070957

van der Kolk, B.A., Roth, S., Pelcovitz, D., Sunday, S., Spinazzola, J., 2005. Disorders of extreme stress: The empirical foundation of a complex adaptation to trauma. J. Trauma. Stress 18, 389–399. 10.1002/jts.20047

Wei, R., Wang, J., Jia, E., Chen, T., Ni, Y., Jia, W., 2018. GSimp: A Gibbs sampler based left-censored missing value imputation approach for metabolomics studies. PLoS Comput. Biol. 14, e1005973. 10.1371/journal.pcbi.1005973

Williams, J.A., Burgess, S., Suckling, J., Lalousis, P.A., Batool, F., Griffiths, S.L., Palmer, E., Karwath, A., Barsky, A., Gkoutos, G.V., Wood, S., Barnes, N.M., David, A.S., Donohoe, G., Neill, J.C., Deakin, B., Khandaker, G.M., Upthegrove, R., PIMS Collaboration, 2022. Inflammation and Brain Structure in Schizophrenia and Other Neuropsychiatric Disorders: A Mendelian Randomization Study. JAMA Psychiatry 79, 498–507. 10.1001/jamapsychiatry.2022.0407

Zung, W.W., Richards, C.B., Short, M.J., 1965. Self-rating depression scale in an outpatient clinic. Further validation of the SDS. Arch. Gen. Psychiatry 13, 508–515. 10.1001/archpsyc.1965.01730060026004

