## Supplementary Figure S1 for "Neuroimmune signatures linking inflammatory proteomics to temporal cortical structure in mothers who perpetrated child maltreatment"

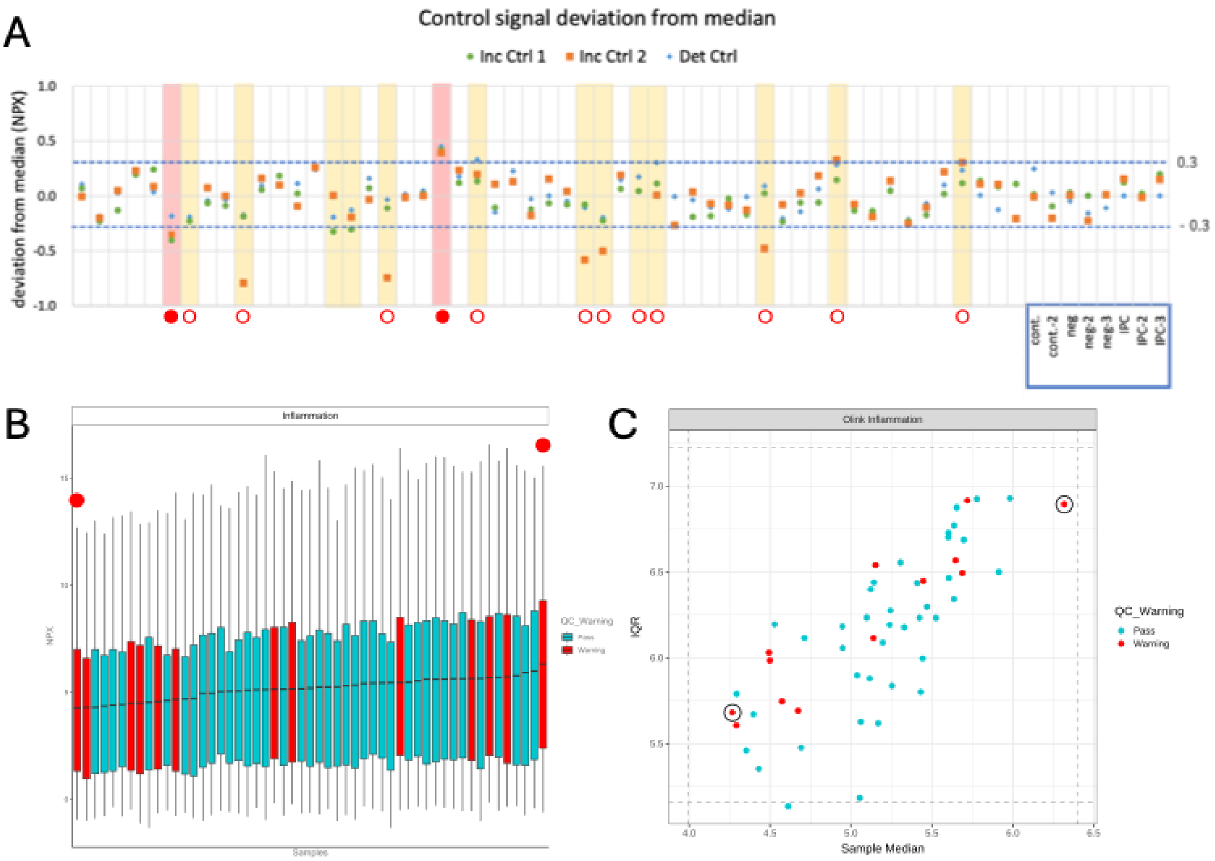


**Supplementary Figure S1. QC assessment and sample outlier identification for the Olink Target 96 Inflammation panel.**

**(A)** Deviation of internal controls (Incubation Control 1 & 2, Detection Control) from the median across all samples. Samples are arrayed along the x-axis. Samples flagged with a QC warning are highlighted. Red-filled circles (●) indicate the two samples with the most prominent deviations, which are further inspected in panels B and C. **(B)** Distribution of Normalized Protein eXpression (NPX) values for all proteins across each sample, ordered by median NPX. Samples with a QC warning are shown in red. The two most prominent outlier samples identified in (A) are marked with large red dots. **(C)** Sample Interquartile Range (IQR) versus median NPX plot. This plot visualizes sample variability against the central tendency. The two outlier samples are highlighted with black circles. Based on this comprehensive assessment, the sample with the highest median NPX (top-right in panel C) was identified as a clear technical outlier due to its significant deviation across all QC metrics and was excluded from subsequent analyses. Although the sample with the lowest median NPX (bottom-left in panel C) was also flagged, it clustered closely with other samples that passed QC. Therefore, it was considered a marginal outlier and was retained in the final dataset.
